# Cervical Stiffness Index as predictor of preterm birth in women with threatened preterm labor

**DOI:** 10.1101/2025.06.09.25328886

**Authors:** Dario Colacurci, Gabriele Saccone, Alessandra Ammendola, Giorgia Buonomo, Chiara Murolo, Mariavittoria Locci

## Abstract

**Objective:** To evaluate the accuracy of Cervical Stiffness Index (CSI) in predicting preterm birth (PTB) in women with threatened preterm labor (PTL).

**Study design:** This was a single-center, prospective, observational study of women with singleton pregnancies presenting to obstetrics triage for threatened PTL between 24^0/7^ and 33^6/7^ weeks. Women included in the study received a physical examination, ultrasound examination, transvaginal ultrasound cervical length measurement, and cervical stiffness assessment.

**Results:** Between April 2022 and August 2024, 100 women with signs and symptoms of PTL were recruited. Thirty-five had a PTB, of which 7 delivered < 34 weeks, 5 within 14 days and 11 within 28 days from measurement. Median CSI and cervical length (CL) were significantly different between women delivering at term and women with a PTB. The Area under the Curve of CSI and CL for prediction of delivery ≤ 14 days from measurement were 0.979 (95% CI, 0.952–1.000) and 0.744 (0.369–1.000), respectively; for delivery ≤ 28 days from measurement, 0.802 (0.612–0.993), and 0.764 (0.579–0.948), respectively; for PTB < 34 weeks, 0.873 (0.729–1.000) and 0.836 (0.563–1.000), respectively; and for PTB < 37 weeks 0.845 (0.763–0.926) and 0.779 (0.680– 0.877), respectively.

**Conclusions:** Women presenting with threatened PTL and ultimately delivering prematurely have a significantly softer cervix than women delivering at term, showing the potential of Cervical Stiffness Index as a predictor of PTB in symptomatic women.

**Clinical trial registration:** NCT05355649

## 1. Introduction

Preterm birth (PTB) is the leading cause of infant mortality, and those who survive could have short- and long-term complications such as respiratory distress necrotizing enterocolitis, retinopathy, bronchopulmonary dysplasia, intraventricular hemorrhage, cognitive impairment and behavioral problems [1]. Despite new technologies and modern medicine, the rate of PTBs is still increasing [2]. The current practice for prevention remains the secondary screening and early diagnosis of women at risk of PTB. The success of this strategy depends on early identification of signs of preterm labor, and on the correct timing of delivering treatments such as tocolytics and steroids [3–10].

So far, the gold standard approach for screening and diagnosis of PTB is based on transvaginal ultrasound cervical length (TVU CL) measurement, and evaluation of clinical signs of preterm labor (PTL) [11–14]. The management of women presenting with PTL remains a challenge as clinicians cannot yet clearly identify women at true risk of PTL and differentiate them from those who will not deliver soon, or even will deliver at term. Current diagnostic methods are insufficient, as over 50% of the women presenting with PTL symptoms do not deliver prematurely [14] but are still unnecessarily hospitalized and treated with corticosteroids and tocolytics with resultant increases in healthcare costs, and only less than 10% of women give birth within 7 days of presentation [15].

In recent years, several trials have evaluated new approaches for early screening of PTB, including use of biomarker tests, such as the fetal fibronectin (fFN) and phosphorylated insulin-like growth factor binding protein-1 (phIGFBP1) [16–19]. Several researchers have started to evaluate the use of a novel aspiration-based device for measuring cervical stiffness, the Pregnolia System, to assess cervical remodeling in several populations [20–28]. However, results from clinical trials in women with threatened PTL have not yet been performed.

The aim of this prospective study was to evaluate the diagnostic accuracy potential of cervical stiffness measured with the Pregnolia System in predicting PTB in women with threatened PTL.

## 2. Materials and methods

This was a single-center, prospective, observational study including women aged 18 years or older, with singleton pregnancies and presenting to obstetrics triage for threatened PTL between 24^0/7^ and 33^6/7^ weeks from April 2022 to August 2024. Threatened PTL was characterized by the presence of one or more of the following symptoms: uterine contractions, abdominal cramps, pelvic pressure, lower back pain, vaginal bleeding [6]. Exclusion criteria were multiple gestations, rupture of membranes, vaginal bleeding that could not be stopped, placenta previa, cerclage or pessary in situ, cervical dilatation ≥ 3 cm, fetal malformations, fetal death, and cervical cancer.

Women were assessed according to an approved clinical guideline based on transvaginal ultrasound cervical length screening [14], recommending intervention for PTL (i.e. admission, tocolysis, and steroids) for women with TVU CL <30 mm, and discharge for women with threatened PTL but with TVU CL ≥30 mm.

The trial was approved by the local ethics committee. All participants in the trial provided written informed consent. The trial protocol was registered on Clinicaltrials.gov before enrollment (https://clinicaltrials.gov/study/NCT05355649).

### 2.1. Measurements of cervical stiffness

Women included in the study received a physical examination, detailed ultrasound examination, TVU CL measurement, and cervical stiffness assessment.

Cervical stiffness was measured by using the CE-marked Pregnolia System (Pregnolia AG, Wiesenstrasse 33, Schlieren, Switzerland) [21,22]. The device probe was placed with the help of a speculum, transvaginally, on the anterior lip of the cervix, at 12 o’clock position (Figure 1). To determine the tissue stiffness, the cervical tissue was slowly pulled into the probe tip. The vacuum level required to displace the tissue into the probe tip by a fixed distance (4 mm) characterizes the tissue stiffness, called Cervical Stiffness Index (CSI), and is expressed in mbar. The stiffer the cervical tissue, the higher the vacuum needed to pull it. Three consecutive measurements were performed, with the probe at the same position and without any time lag.

**Figure 1:**
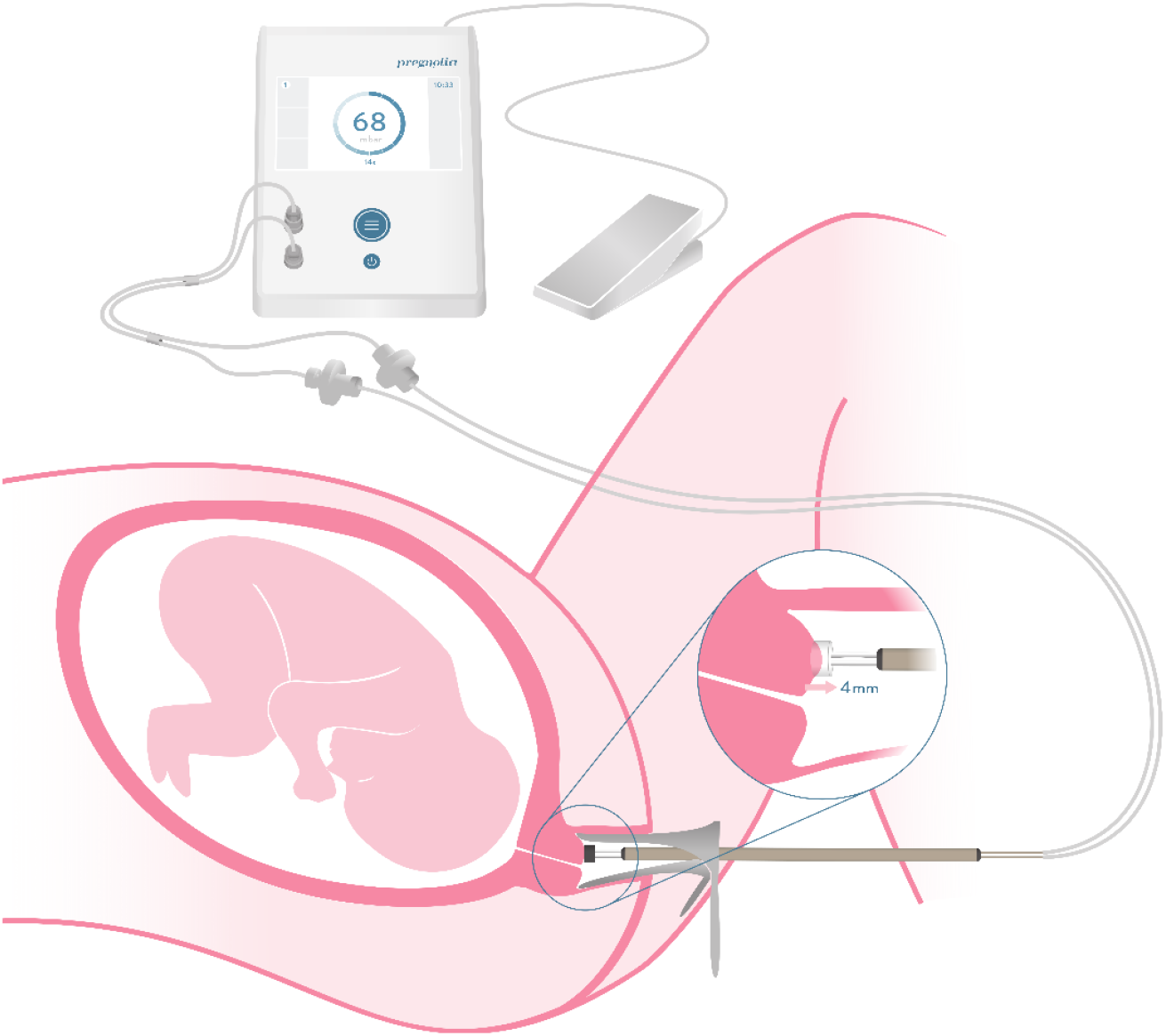
Pregnolia System overview. General overview of the Pregnolia System and its application. The probe is manually placed on the cervix during a routine gynecological evaluation, with the aid of a speculum. Figure adapted with kind permission of Pregnolia AG.

### 2.2. Study objectives

The study primary objective was to determine the value of CSI in women presenting with symptoms of PTL as measured by the Pregnolia System.

The study further aimed at determining the correlation of CSI with pregnancy outcomes (gestational age at birth, time to delivery) and determining if the detection of true PTL and of false PTL improves when CSI is added to the standard screening method (TVU CL).

### 2.3. Statistical analysis

Three repeated CSI measurements were obtained sequentially, and the median value was used in the analyses. Results for each parameter are reported as median (interquartile range [IQR], using R quantile function, type 7), unless otherwise specified.

CSI values are not normally distributed, but lognormally. To determine statistical significance and p-values, we used, as appropriate: the Wilcoxon Rank Sum Test for comparisons between two groups on continuous variables, Kruskal–Wallis test for comparisons among more than two groups on continuous variables, Fisher’s Exact Test for dichotomous categorical variables, and Pearson’s Chi-squared Test to assess the independence between groups on categorical variables with more than two values. Probability density distributions were calculated by fitting a lognormal distribution using the maximum-likelihood method. A p-value of < 0.05 was considered significant. All analysis was performed in R (R Core Team 2024, [29]) using the R stats package for statistical significance determination and model evaluation.

The combined performance of CSI and CL was assessed using a linear model to predict gestational age (GA) (before <37 weeks’ and <34 weeks’ gestation, and within ≤14 days and ≤ 28 days from measurement - *model GA*) or time to delivery (TTD) within ≤14 days and ≤ 28 days from measurement (*model TTD*). This analysis employed the lm function from R’s stats library. GA at measurement and smoking status were initially included in the model as potential confounding factors and assessed for significance. Variables that were not statistically significant (p > 0.05) and were not identified as confounding factors were excluded from the final model.

Predictive performance for PTB < 37 and < 34 weeks and delivery ≤ 14 and ≤ 28 days were calculated as area under receiver-operating-characteristics curve (AUC) for CSI and CL alone, and in combination by using linear models. DeLong’s test was used to assess for a difference in the AUC of the receiver-operating-characteristics (ROC) curves. The DeLong’s test was performed by using the roc.test function from R’s pROC package. Thresholds for each predictor were selected in order to maximize the sensitivity for at least 95% specificity; in addition, a threshold of 25 mm for cervical length was also selected, as it is a commonly used cutoff for PTB prediction [30].

A sample size of 100 participants was planned *a priori*.

## 3. Results

During the study period, 100 women with singleton pregnancies and threatened PTL between 24 and 34 weeks of gestation consented to take part in the study and were enrolled. Table 1 shows the baseline demographic characteristics for enrolled participants.

**Table 1.**
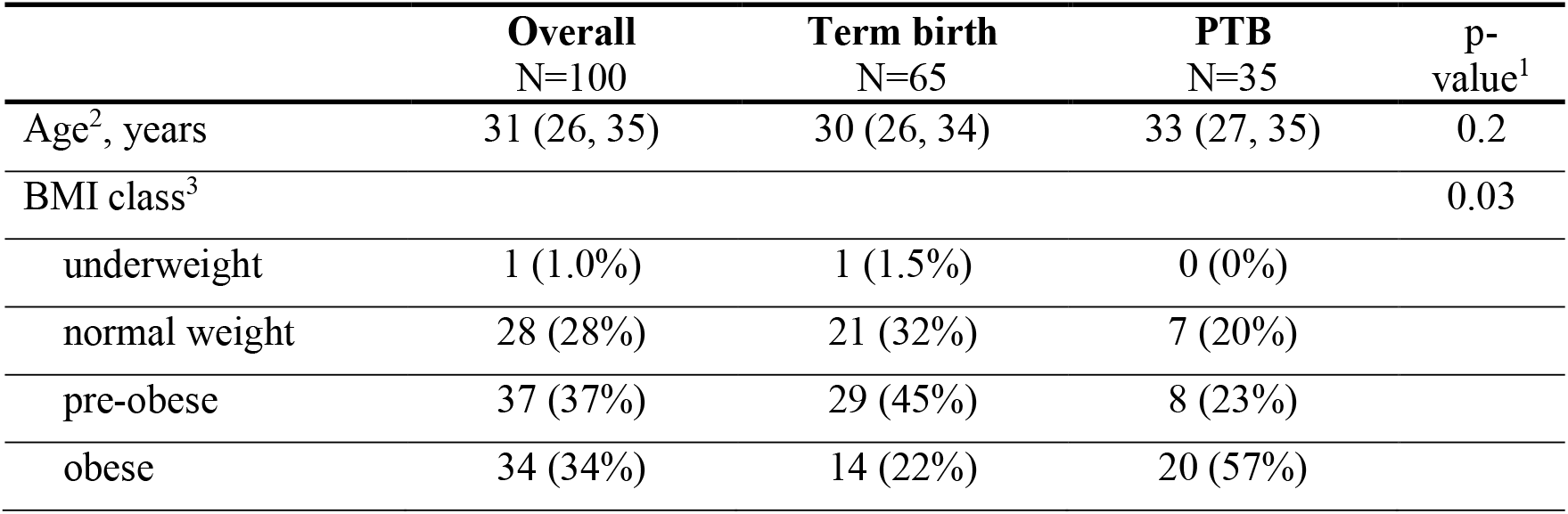
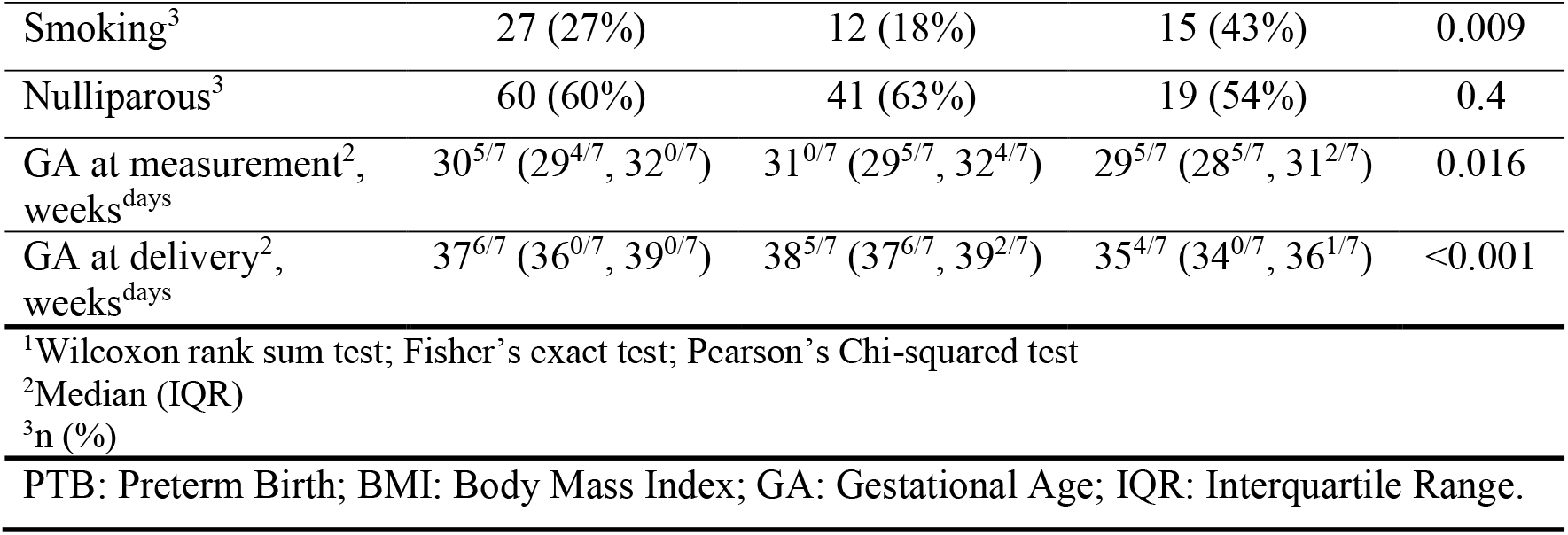
Baseline demographic characteristics for the study population.

|  | Overall<br>N=100 | Term birth<br>N=65 | PTB<br>N=35 | p-value<sup>1</sup> |
| --- | --- | --- | --- | --- |
| Age<sup>2</sup>, years | 31 (26, 35) | 30 (26, 34) | 33 (27, 35) | 0.2 |
| BMI class<sup>3</sup> |  |  |  | 0.03 |
| underweight | 1 (1.0%) | 1 (1.5%) | 0 (0%) |  |
| normal weight | 28 (28%) | 21 (32%) | 7 (20%) |  |
| pre-obese | 37 (37%) | 29 (45%) | 8 (23%) |  |
| obese | 34 (34%) | 14 (22%) | 20 (57%) |  |
| Smoking<sup>3</sup> | 27 (27%) | 12 (18%) | 15 (43%) | 0.009 |
| Nulliparous<sup>3</sup> | 60 (60%) | 41 (63%) | 19 (54%) | 0.4 |
| GA at measurement<sup>2</sup>,<br>weeks<sup>days</sup> | 30<sup>5/7</sup> (29<sup>4/7</sup>, 32<sup>0/7</sup>) | 31<sup>0/7</sup> (29<sup>5/7</sup>, 32<sup>4/7</sup>) | 29<sup>5/7</sup> (28<sup>5/7</sup>, 31<sup>2/7</sup>) | 0.016 |
| GA at delivery<sup>2</sup>,<br>weeks<sup>days</sup> | 37<sup>6/7</sup> (36<sup>0/7</sup>, 39<sup>0/7</sup>) | 38<sup>5/7</sup> (37<sup>6/7</sup>, 39<sup>2/7</sup>) | 35<sup>4/7</sup> (34<sup>0/7</sup>, 36<sup>1/7</sup>) | <0.001 |
| <sup>1</sup>Wilcoxon rank sum test; Fisher's exact test; Pearson's Chi-squared test |  |  |  |  |
| <sup>2</sup>Median (IQR) |  |  |  |  |
| <sup>3</sup>n (%) |  |  |  |  |
| PTB: Preterm Birth; BMI: Body Mass Index; GA: Gestational Age; IQR: Interquartile Range. |  |  |  |  |

Of the 100 enrolled women, 65 (65%) delivered at term and 35 (35%) had a PTB < 37 weeks, of which 7 delivered < 34 weeks (7%) (Table 2), and 5 (5%) ≤ 14 days and 11 (11%) ≤ 28 days from the measurement (Table 3). One woman (1%) delivered within 1 week from the measurement.

**Table 2.** Median CSI (mbar) and CL (mm) for birth outcome.

|  | **Term**<br>**birth**<br>**N=65** | **PTB**<br>**N=35** | **p-**<br>**value<sup>1</sup>** | **Term**<br>**birth**<br>**N=65** | **Late PTB**<br>**N=28** | **Early PTB**<br>**N=7** | **p-**<br>**value<sup>2</sup>** |
| --- | --- | --- | --- | --- | --- | --- | --- |
| Median<br>CSI<br>(mbar) | 88<br>(70, 102) | 50<br>(36, 64) | **<0.001** | 88<br>(70, 102) | 52<br>(44, 66) | 20<br>(18, 41) | **<0.001** |
| CL (mm) | 29.0<br>(24.0, 31.0) | 22.0<br>(18.5, 24.0) | **<0.001** | 29.0<br>(24.0, 31.0) | 23.0<br>(20.8, 24.5) | 15.0<br>(7.5, 17.5) | **<0.001** |

<sup>1</sup>Wilcoxon rank sum test
<sup>2</sup>Kruskal-Wallis rank sum test
PTB: Preterm Birth; CSI: Cervical Stiffness Index; CL: Cervical Length.
Values are reported as Median (interquartile range). Early PTB is defined as a birth <34 weeks, late PTB as a birth ≥ 34 and < 37 weeks.

**Table 3.**
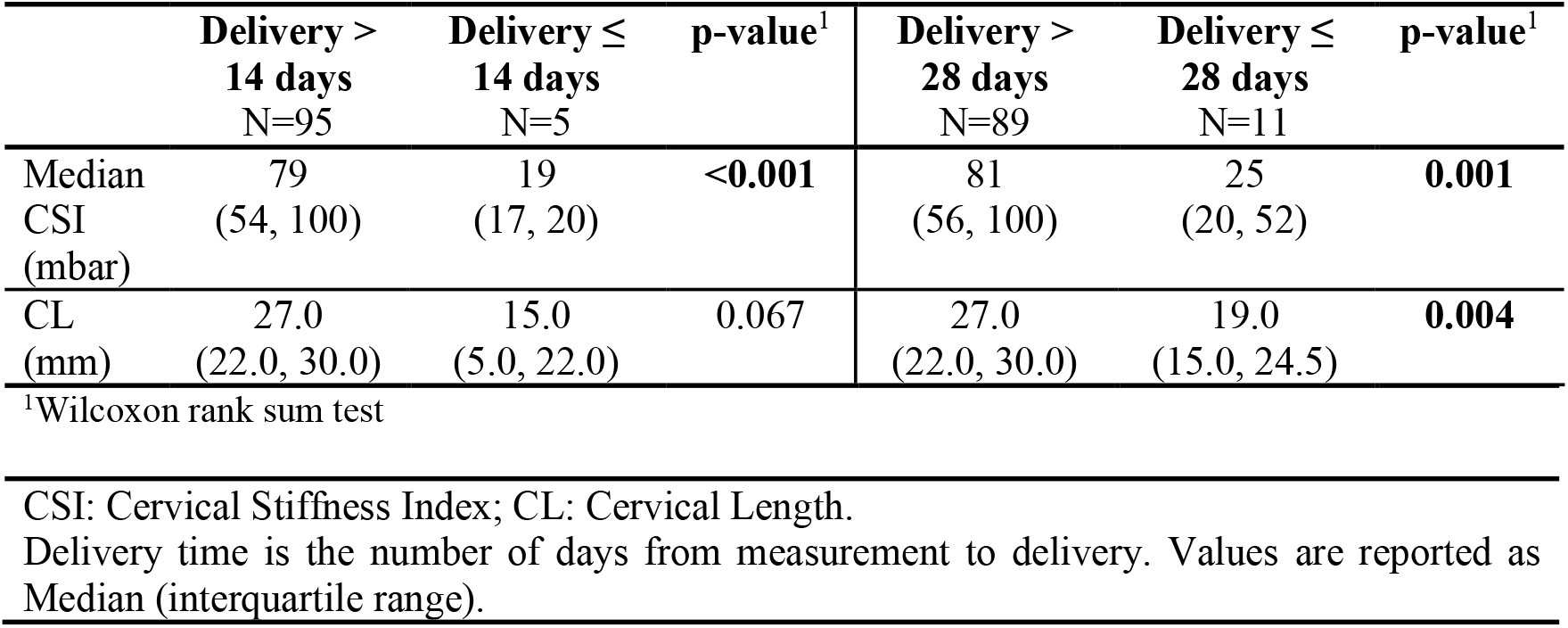
Median CSI (mbar) and CL (mm) for time to delivery.

Symptomatic women who delivered preterm had a significantly softer and shorter cervix compared to those who delivered at term (Table 2). Also, women who delivered within 14 and 28 days from measurement (i.e. from admission for PTL), had a significantly softer cervix. The TVU CL was shorter, but reached significance only for delivery ≤ 28 days (Table 3).

CSI showed a non-significant trend towards improved predictive performance compared to CL to predict PTB < 37 weeks (AUC 0.845 (95% CI, 0.763–0.926) for CSI vs 0.779 (0.680–0.877) for CL); PTB < 34 weeks (AUC 0.873 (0.729–1.000) for CSI vs 0.836 (0.563-1.000) for CL), delivery ≤ 14 days from measurement (AUC 0.979 (0.952–1.000) for CSI vs 0.744 (0.369–1.000) for CL), and delivery ≤ 28 days from measurement (AUC 0.802 (0.612–0.993) for CSI vs 0.764 (0.579 0.948) for CL) (Figure 2).

**Figure 2:**
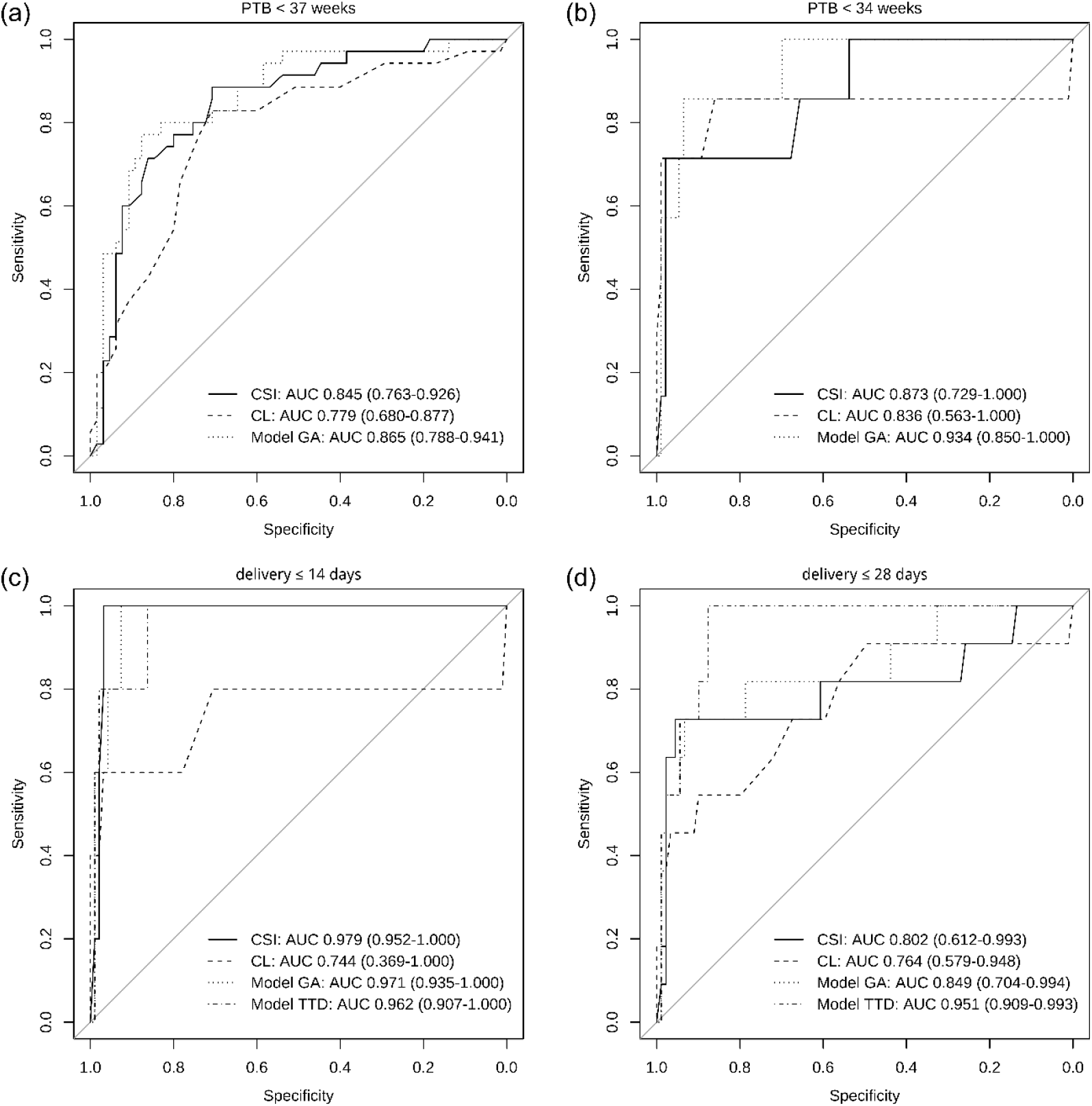
Receiver-operating-characteristics curves. Receiver-operating-characteristics (ROC) curves showing predictive accuracy of Cervical Stiffness Index (CSI), cervical length (CL), the linear model to predict gestational age (model GA) and the linear model to predict time to delivery (model TTD) for (a) preterm birth (PTB) <37 weeks’ gestation, (b) PTB < 34 weeks’ gestation, (c) delivery ≤ 14 days from measurement and (d) delivery ≤ 28 days from measurement. Area under the ROC curves (AUCs) with 95% Confidence Interval are reported in the plots.

The final models to predict GA and TTD included log of median CSI and CL as independent variables and excluded smoking, which was non-significant and did not contribute to improving the model’s performance. GA at measurement was a significant variable in the *model TTD* and thus was included in that model only.

The combination of CSI and CL in the linear models generally enhanced the AUC (Figure 2). The improvement in AUC was significant when comparing the prediction of PTB < 37 weeks using CL alone to the combination of CSI and CL in the model (p=0.030 with a DeLong’s test for two correlated ROC curves). The improvement was also significant for predicting delivery ≤ 28 days from measurement when comparing CL alone to the linear model for predicting TTD (p=0.038), indicating that in these cases adding CSI to CL measurement significantly improves the prediction performance.

Predictive performances in terms of sensitivity (SN), specificity (SP), positive predictive value (PPV) and negative predictive value (NPV) for predicting PTB < 37 and < 34 weeks and delivery ≤ 14 and ≤ 28 days from measurement are reported in Table 4.

**Table 4.** Sensitivity (SN), specificity (SN), positive and negative predictive values (PPV and NPV) for predicting PTB < 37 and < 34 weeks and delivery ≤ 14 and ≤ 28 days from measurement.

| PTB < 37 weeks (n=35) |  |  |  |  |  |
| --- | --- | --- | --- | --- | --- |
|  | Threshold | SN | SP | NPV | PPV |
| CSI | < 39 mbar | 28.6%<br>(13.6, 43.5) | 95.4%<br>(90.3, 100) | 76.9%<br>(54, 99.8) | 71.3%<br>(61.8, 80.8) |
| CL | ≤ 16.5 mm | 20%<br>(6.75, 33.3) | 98.5%<br>(95.5, 100) | 87.5%<br>(64.6, 100) | 69.6%<br>(60.2, 79) |
| CL | ≤ 25 mm | 77.1%<br>(63.2, 91.1) | 73.8%<br>(63.2, 84.5) | 61.4%<br>(47, 75.8) | 85.7%<br>(76.5, 94.9) |
| Model GA | ≤ 36.580 | 48.6%<br>(32, 65.1) | 95.4%<br>(90.3, 100) | 85%<br>(69.4, 100) | 77.5%<br>(68.3, 86.7) |
| PTB < 34 weeks (n=7) |  |  |  |  |  |
|  | Threshold | SN | SP | NPV | PPV |
| CSI | < 23 mbar | 71.4%<br>(38, 100) | 97.8%<br>(94.9, 100) | 71.4%<br>(38, 100) | 97.8%<br>(94.9, 100) |
| CL | ≤ 15.5 mm | 71.4%<br>(38, 100) | 98.9%<br>(96.8, 100) | 83.3%<br>(53.5, 100) | 97.9%<br>(95, 100) |
| CL | ≤ 25 mm | 85.7%<br>(59.8, 100) | 59.1%<br>(49.1, 69.1) | 13.6%<br>(3.5, 23.8) | 98.2%<br>(94.7, 100) |
| Model GA | ≤ 34.956 | 57.1%<br>(20.5, 93.8) | 98.9%<br>(96.8, 100) | 80%<br>(44.9, 100) | 96.8%<br>(93.3, 100) |
| Delivery ≤ 14 days (n=5) |  |  |  |  |  |
|  | Threshold | SN | SP | NPV | PPV |
| CSI | < 26 mbar | 100%<br>(100, 100) | 96.8%<br>(93.3, 100) | 62.5%<br>(29, 96) | 100%<br>(100, 100) |
| CL | ≤ 15.5 mm | 60%<br>(17.1, 100) | 96.8%<br>(93.3, 100) | 50%<br>(9.99, 90) | 97.9%<br>(95, 100) |
| CL | ≤ 25 mm | 80%<br>(44.9, 100) | 57.9%<br>(48, 67.8) | 9.1%<br>(0.596, 17.6) | 98.2%<br>(94.7, 100) |
| Model GA | ≤ 35.663 | 80%<br>(44.9, 100) | 95.8%<br>(91.8, 99.8) | 50%<br>(15.4, 84.6) | 98.9%<br>(96.8, 100) |
| Model TTD | ≤ 3.359 | 80%<br>(44.9, 100) | 97.9%<br>(95, 100) | 66.7%<br>(28.9, 100) | 98.9%<br>(96.9, 100) |
| Delivery ≤ 28 days (n=11) |  |  |  |  |  |
|  | Threshold | SN | SP | NPV | PPV |
| CSI | < 37 mbar | 72.7%<br>(46.4, 99) | 95.5%<br>(91.2, 99.8) | 66.7%<br>(40, 93.3) | 96.6%<br>(92.8, 100) |
| CL | ≤ 16.5 mm | 45.5%<br>(16, 74.9) | 96.6%<br>(92.9, 100) | 62.5%<br>(29, 96) | 93.5%<br>(88.4, 98.5) |
| **CL** | $\leq 25$ mm | 72.7%<br>(46.4, 99) | 59.6%<br>(49.4, 69.7) | 18.2%<br>(6.79, 29.6) | 94.6%<br>(88.7, 100) |
| **Model GA** | $\leq 35.663$ | 54.5%<br>(25.1, 84) | 97.8%<br>(94.7, 100) | 75%<br>(45, 100) | 94.6%<br>(89.9, 99.2) |
| **Model TTD** | $\leq 3.772$ | 54.5%<br>(25.1, 84) | 97.8%<br>(94.7, 100) | 75%<br>(45, 100) | 94.6%<br>(89.9, 99.2) |
| PTB: Preterm Birth; CSI: Cervical Stiffness Index; CL: Cervical Length; GA: Gestational Age; TTD: Time to Delivery.<br>Model GA: linear model to predict gestational age. Model TTD: linear model to predict time to delivery. Values are reported as % (95% Confidence Interval). |  |  |  |  |  |

## 4. Discussion

This single-center study aimed to evaluate the performance of Cervical Stiffness Index measured with the Pregnolia System to predict PTB in women with a singleton pregnancy presenting with threatened PTL. One hundred women presenting to obstetrics triage for threatened PTL between 24^0/7^ and 33^6/7^ weeks of gestation were included. Our results have shown that cervical stiffness in women with a PTB is significantly softer than in women with a term birth, and that cervical stiffness has a good predictive performance for delivery within 14 and 28 days and prediction of PTB < 34 and < 37 weeks.

The strength of our study is that, to our knowledge, this is the first study investigating differences in cervical stiffness measured with an aspiration-based device between women with threatened PTL who ultimately delivered prematurely and those who delivered at term. The Pregnolia System has been, or is currently being, evaluated in other studies and in different populations [20–28], however this is the first time that data on women with threatened PTL are reported. As the fetal fibronectin test has been withdrawn by its manufacturer from the European market [31], it is relevant to evaluate new tests and biomarkers that can contribute to a better diagnosis of women at risk of imminent delivery. Our study is contributing towards the evaluation of new PTB predictors and providing clinical evidence related to their performance.

In addition, the data were collected prospectively in a single center, thus making the data collection uniform.

The main limitation is that the study was explorative and, given the lack of previous data, could not be powered to formally investigate our hypothesis. Nevertheless, in our cohort we had 35 women delivering prematurely, a reasonable number to draw meaningful preliminary conclusions. However, we could not analyze prediction of delivery within 7 days – a most relevant clinical outcome for informing patient management – as only one woman delivered within this time period. The low rate of women delivering within 7 days can potentially be explained by over diagnosis, given the broad symptoms used in this study and in our unit to evaluate threatened preterm labor.

Novel tools such as the Pregnolia System, and methods like the assessment of cervical maturation, may improve the identification of women at true risk of imminent labor and thus aid management of this vulnerable population. Our results demonstrate value in assessing cervical stiffness as a parameter used either alone or in combination with the measurement of cervical length. However, our results still need to be replicated in a larger cohort and reproduced by other groups to prove robustness.

## 5. Conclusion

Our study has shown that women presenting with threatened PTL and ultimately delivering prematurely have a significantly softer cervix than women delivering at term.

Cervical Stiffness Index measured with the Pregnolia System has potential to identify women at true risk of PTL. However, further evaluation and studies are needed to demonstrate robustness of data for the use of CSI as a predictor of preterm birth and in the management of these patients.

## Data Availability

All data produced in the present study are available upon reasonable request to the authors

